# Accelerated epigenetic aging and prospective morbidity and mortality among U.S. veterans

**DOI:** 10.1101/2024.10.23.24315691

**Authors:** Kyle J. Bourassa, Livia Anderson, Sandra Woolson, Paul A. Dennis, Melanie E. Garrett, Lauren Hair, Michelle Dennis, Karen Sugden, Benjamin Williams, Renate Houts, VA Mid Atlantic MIRECC Workgroup, Patrick S. Calhoun, Jennifer C. Naylor, Allison E. Ashley-Koch, Jean C. Beckham, Avshalom Caspi, Gregory A. Taylor, Katherine S. Hall, Terrie E. Moffitt, Nathan A. Kimbrel

## Abstract

Epigenetic measures of aging derived from DNA methylation are promising biomarkers associated with prospective morbidity and mortality, but require validation in real-world medical settings. Using data from 2,216 post-9/11 veterans, we examined whether accelerated DunedinPACE aging scores were associated with chronic disease morbidity, predicted healthcare costs, and mortality assessed over an average of 13.1 years of follow up in VA electronic health records. Veterans with faster DunedinPACE aging scores developed more chronic disease and showed larger increases in predicted healthcare costs over the subsequent 5, 10, and 15 years. Faster aging was associated with incident myocardial infarction, stroke, diabetes, cancer, liver disease, and renal disease, as well greater risk of mortality due to all-causes and chronic disease. These findings provide evidence that accelerated epigenetic aging is associated with worsening prospective health across multiple chronic diseases and organ systems assessed using electronic health records from an integrated healthcare system.

---

Epigenetic measures of aging derived from DNA methylation (DNAm) developed over the last decade can assess biological aging using biological samples collected at a single point in time^1-5^. This new development has the potential to allow for the identification of individuals with accelerated aging who could be targeted by geroprotective interventions. If epigenetic measures of aging are associated with prospective health, then slowing the rate at which individuals are aging, defined in geroscience theory as a common cause of chronic disease morbidity and mortality^6-7^, would be expected to improve health across many chronic disease pathways and organ systems^9-11^. Aging biomarkers would be invaluable surrogate health outcomes randomized control trials testing interventions that aim to slow aging and improve health^12^, as well as observational studies investigating health trajectories^13^. With additional validation and refinement, such measures also have the potential to serve as clinical biomarkers of future health for use in clinical settings.

There is promising evidence that second- and third-generation epigenetic measures of aging are associated with future health^1-2,14-15^, particularly third-generation measures developed using longitudinal biomarker data^1,15^ (e.g., DunedinPACE^2^). However, realizing the potential clinical and research value of epigenetic measures of aging requires evaluating whether these biomarkers are associated with prospective chronic disease morbidity and mortality in real-world medical settings. To do so, we integrated survey, epigenetic, and electronic health record (EHR) data from 2,216 U.S. military veterans who served after September 11, 2001^16^. The cohort was 37.4 years old at enrollment with an average follow-up of 13.1 years, which afforded an observation window spanning early adulthood into midlife and older age, periods that often include the onset of chronic disease^17^.

## Methods

### Participants and Study Design

Participants were enrolled in the VISN 6 Mid-Atlantic Mental Illness Research, Education, and Clinical Center’s (MIRECC’s) Post-Deployment Mental Health Study (PDMH^16^), a multi-site study of veterans who served in the post-9/11 period. The Durham, Richmond, and Salisbury VA Medical Centers’ Institutional Review Boards approved the PDMH study protocol and all participants provided informed consent. The study included participants with DNAm and VA EHR data (**eFigure 1**), resulting in a sample of 2,216 veterans followed an average of 13.1 years (*SD* = 2.8).

### Measures

#### Demographics

Participants reported their age, sex, race, ethnicity, years of education, and smoking status (coded: never smoker, 0; past smoker, 1; current smoker, 2). Sex, race, and ethnicity self-reports were confirmed using sex chromosomes and ancestry values derived from genetic data.

#### Genomic DNAm data generation and processing

Whole blood was collected during baseline assessments and analyzed using the Infinium HumanMethylation450 or MethylationEPIC Beadchip (Illumina Inc., San Diego, CA) to derive DNAm data^18-19^. Internal replicates were included and checked for consistency using single nucleotide polymorphisms on each array. Quality control was performed using the minfi^20^ and ChAMP^21^ R packages. Probe quality control and data normalization were performed within each batch using the R package wateRmelon^22^. Raw beta values were normalized using the dasen approach^23^ and batch and chip adjustments were completed using ComBat in the R package sva^24^.

##### DunedinPACE

Epigenetic aging was assessed by applying the DunedinPACE algorithm to PDMH DNAm data^20-21^. The algorithm^25^ is derived from reliable CpG probes^26^ and produces aging scores that represent years of biological aging per chronological year. Statistical tests used continuous DunedinPACE scores unless otherwise noted (four aging quartiles were created for visualization and interpretation).

##### Technical covariates

A dummy variable was created to denote if DNAm data was generated using 450k or EPIC V1 chips. Additional covariates were derived using FlowSorted.Blood.450k and FlowSorted.Blood.EPIC packages to estimate cell count proportions^27^ for white blood cells categories (T lymphocytes (CD4+ and CD8+), B cells (CD19+), monocytes (CD14+), NK cells (CD56+) and Neutrophils).

##### DNAm smoking

Lifetime exposure to tobacco smoke^28^ was calculated for participants using a DNAm measure^29^. Methylation smoking scores correlated with self-reported smoking (*r* = 0.55, *p* < .001).

##### PC-adjusted second-generation epigenetic clocks: PhenoAge and GrimAge

We derived two principle components (PC) adjusted second-generation epigenetic clocks, PC-PhenoAge^30^ and PC-GrimAge^2^. The PhenoAge^30^ algorithm was trained on clinical data from a large training dataset^30^, whereas GrimAge was trained on all-cause mortality data. We derived PC-PhenoAge and PC-GrimAge values using established algorithms^12^ that account for PCs to improve the reliability of clock estimates^12^. Both measures were residualized on chronological age to provide a measure of age acceleration.

#### Electronic health record data

Prospective health outcomes and clinical biomarkers were derived using the VA EHR. **eMethod 1** provides a detailed description of EHR data processing. Veterans enrolled in the PDMH from 2005 to 2016^13^, which resulted in follow-up periods ranging from 7.3 to 18.5 years (88 to 222 months). EHR data coverage (**eTable 1**) was predominantly based on the timing of veterans’ baseline assessment (and resulting length of EHR follow-up; **eFigure 2**). Year of baseline assessment was not associated with DunedinPACE (**eFigure 3**).

##### Charlson comorbidity index

Charlson comorbidity index scores^31^ (CCI) assessed chronic disease burden and were derived using diagnostic ICD-9 and ICD-10 codes^32^ ascertained from outpatient, inpatient, and purchased care data (i.e., community care referrals from VA providers and/or paid by VA sources). Baseline values were calculated on the date of enrollment in the PDMH and were updated for each follow-up period. CCI scores were used to calculate 10-year CCI-predicted mortality risk.

##### Nosos risk adjustment score

Nosos risk adjustment scores^33-34^ represent predicted annual healthcare costs for VA patients based on the Centers for Medicare and Medicaid Hierarchical Condition Categories risk adjustment model. This algorithm was updated to include items specific to the VA, such as priority status and computed costs^35^. Nosos scores are normalized to a mean of 1.0, such that greater values represent higher predicted patient costs (e.g., a score of 1.25 equals 25% higher predicted costs).

#### Chronic disease onset

Chronic disease onset for each of the disease categories in the CCI were ascertained at baseline, as well as at follow-up to a censor date of 12/31/2023. Combined diagnosis data were used to create a measure of time to first chronic disease onset across categories.

#### Clinical biomarkers

Clinical biomarkers included body mass, blood pressure (BP), and heart rate (HR) assessed during VA clinical encounters. Body mass was calculated using the standard formula via height and weight. BP was assessed using systolic BP. HR was assessed using pulse. All measures were assessed at baseline using a 2-year lookback period, producing data for over 75% of veterans who had clinical encounters with these biomarkers assessed (**eTable 1**).

#### Mortality

Dates of death and all-cause mortality status were ascertained using the VA EHR. Months from PDMH baseline to date of death defined time to mortality to a censor date of 1/21/2024. In total, 92 deaths were observed over the follow-up. National Death Index (NDI) data included in the VA Mortality Data Repository^35^ (MDR) provided primary cause of death, which was used to classify deaths as related to acute causes or chronic disease. A subset of recent deaths did not have cause of death data (*n* = 22), as MDR data is currently censored to 12/31/2021^34^. Further excluding acute mortality events (*n* = 30; due to overdoses, accidents, death by suicide, infection, and homicide) left 40 deaths due to chronic diseases, largely cardiovascular diseases (*n* = 17) and cancer (*n* = 15).

### Data Analysis

We tested the association of DunedinPACE epigenetic aging scores with CCI and Nosos scores at baseline, then with change to 5-, 10- and 15-year follow-ups. Models of 5-, 10-, and 15-year change in CCI and Nosos scores controlled for baseline CCI and Nosos scores, respectively. Analyses of CCI scores used zero-inflated Poisson regression models to account for CCI distributions (see **eFigure 4**) and analyses of Nosos scores used linear regression models. We next tested the association between aging scores and incident onset of chronic diseases and mortality using Cox proportional-hazard models. Finally, we conducted two additional sets of analyses to complement the primary results: 1.) stratifying by sex, then race and ethnicity, and 2.) providing results for two PC-adjusted second-generation epigenetic measures of aging. For each set of models, we report estimates controlling for demographic (age, sex, race and ethnicity, and years of education) and DNAm technical covariates (chip type, white blood cell counts), then results controlling for clinical biomarkers (body mass, BP, HR), and two measures of smoking (self-reported smoking and DNAm-derived tobacco smoke exposure). Tables also include bivariate associations controlling for age. Poisson regression models used Monte Carlo simulation to account for missing data, linear regression models used full maximum likelihood estimation (full results excluding any participants with missing data are reported in **eTable 2**), whereas Cox proportional-hazard models included only participants with full data. Inspection of Schoenfeld residual plots and estimates of the interaction of time with aging scores suggest survival curves met the proportional hazard assumption. All model estimates were scaled to 1 *SD* DunedinPACE aging score. Models were run in MPLUS version 8.3^36^ using two-tailed tests with an *a priori* significance level of 0.05.

## Results

The 2,216 veterans (472 women, 1,744 men) included 1,077 non-Hispanic Black and 1,139 non-Hispanic White veterans, with a mean age of 37.4 years (*SD* = 10.1) at baseline.

### Accelerated Aging and Chronic Disease Burden

Veterans with faster DunedinPACE aging scores had greater chronic disease burden at baseline (β, 0.23; 95% CI, 0.10-0.35; *p* < .001). Veterans with faster aging scores developed greater chronic diseases burden over the subsequent 5 years (β, 0.22; 95% CI, 0.13-0.31; *p* < .001), 10 years (β, 0.24; 95% CI, 0.16-0.31; *p* < .001), and 15 years (β, 0.31; 95% CI, 0.20-0.36; *p* < .001). These associations represented 25% (*RR,* 1.25; 95% CI, 14-36%), 27% (*RR,* 1.27; 95% CI, 17-41%), and 36% (*RR,* 1.36; 95% CI, 95% CI, 22-52%) greater relative risk, respectively. Results remained when controlling for clinical biomarkers and smoking measures (**Table 1**). The size of the associations increased as follow-up periods increased in length. At the 5-, 10-, and 15-year follow-ups, the fastest aging veterans had 0.41, 0.92, and 1.84 higher CCI scores than the slowest aging veterans (**Figure 1; eFigure 5**), corresponding to 5.3, 6.2, and 12.0 times greater relative—and 1.3%, 4.7% and 16.5% greater absolute—increase in 10-year CCI-predicted mortality risk, respectively (**eTable 3**).

**Figure 1.**
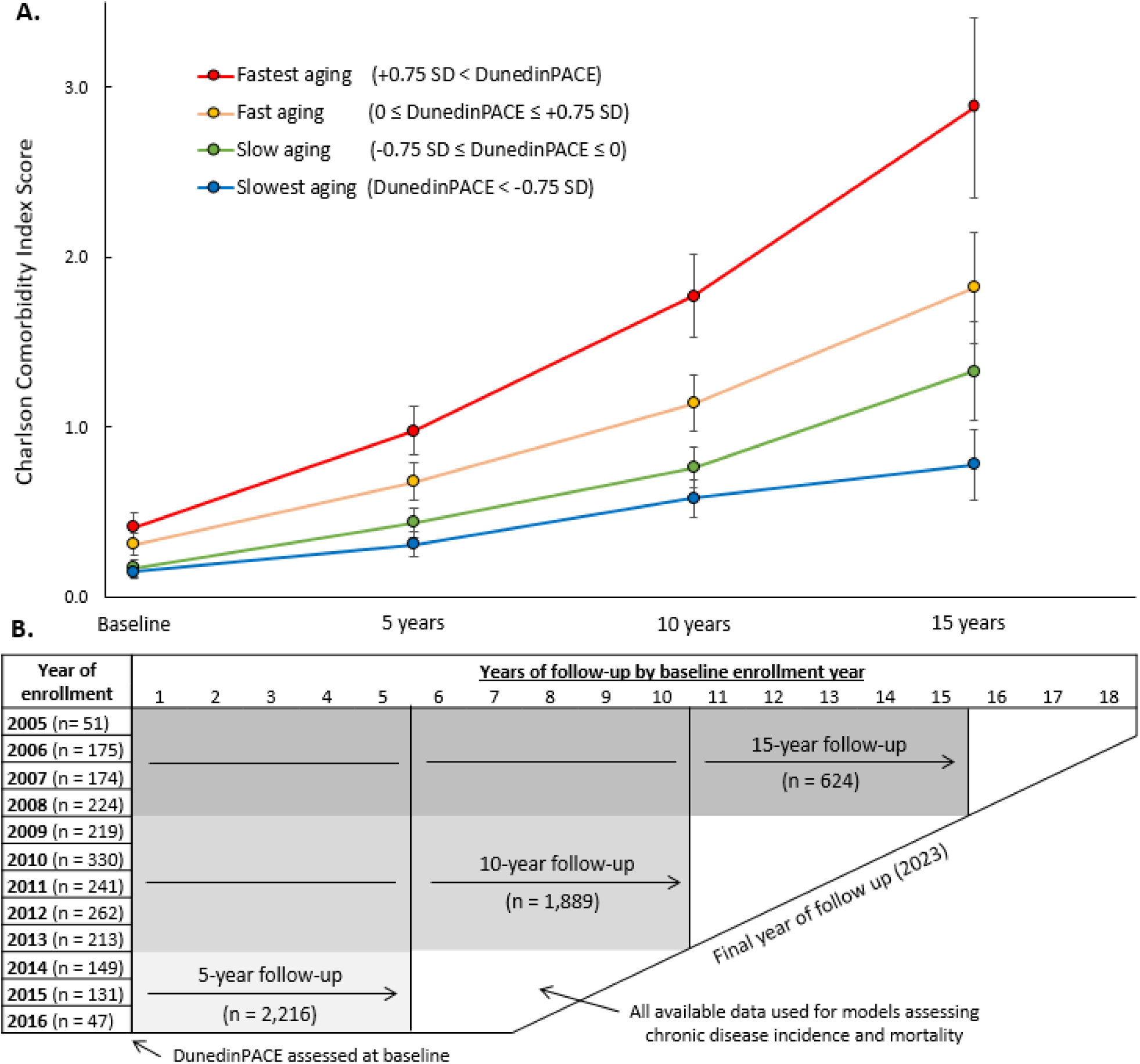
Panel A presents mean CCI scores over time grouped by DunedinPACE aging scores. Four groups were created by standardizing DunedinPACE scores and creating cutoffs at the mean and 0.75 *SD* above and below the mean, corresponding to DunedinPACE values of “slowest aging” ≤ 0.98 (*n* = 517, 23.3%), “slow aging” between 0.98 to 1.07 (*n* = 656, 29.6%), “fast aging” between 1.07 to 1.15 (*n* = 561, 25.3%), and 1.15 ≤ for “fastest aging” (*n* = 482, 21.8%). Groups were created using *a priori* SD cutoffs to roughly approximate quartlies for illustrative purposes—all model estimates used full DunedinPACE aging scores. Panel B presents the study sample by baseline year of enrollment in the PDMH and years of follow-up to the censor date. Baseline PDMH enrollment included the blood draw used to derive DNA methylation data from whole blood.

**Table 1.** Association of DunedinPACE and prospective health among post-9/11 veterans.

| Association with<br>DunedinPACE | Age-adjusted<br>bivariate association |  | Adding demographic and<br>technical covariates |  | Adding clinical<br>biomarkers |  | Adding self-reported and<br>methylation smoking |  |
| --- | --- | --- | --- | --- | --- | --- | --- | --- |
| *N* = 2,216 | $\beta$ | 95% CI | $\beta$ | 95% CI | $\beta$ | 95% CI | $\beta$ | 95% CI |
| **Chronic disease burden (Charlson comorbidity index score)** |  |  |  |  |  |  |  |  |
| Baseline CCI score | 0.25** | [0.14, 0.36] | 0.23** | [0.10, 0.35] | 0.20** | [0.07, 0.33] | 0.19* | [0.01, 0.36] |
| 5-year change in CCI | 0.20** | [0.13, 0.28] | 0.22** | [0.13, 0.31] | 0.17** | [0.07, 0.26] | 0.14** | [0.04, 0.25] |
| 10-year change in CCI | 0.19** | [0.12, 0.26] | 0.24** | [0.16, 0.31] | 0.19** | [0.11, 0.27] | 0.18** | [0.10, 0.26] |
| 15-year change in CCI | 0.29** | [0.19, 0.38] | 0.31** | [0.20, 0.42] | 0.27** | [0.16, 0.38] | 0.30** | [0.18, 0.42] |
| **Predicted annual VA healthcare costs (Nosos risk adjustment score)** |  |  |  |  |  |  |  |  |
| Baseline Nosos score | 0.09** | [0.04, 0.12] | 0.11** | [0.06, 0.15] | 0.11** | [0.03, 0.10] | 0.04 | [-0.01, 0.09] |
| 5-year change in Nosos | 0.09** | [0.05, 0.14] | 0.08** | [0.03, 0.13] | 0.07** | [0.01, 0.12] | 0.03 | [-0.03, 0.10] |
| 10-year change in Nosos | 0.24** | [0.17, 0.31] | 0.23** | [0.15, 0.31] | 0.23** | [0.14, 0.31] | 0.22** | [0.12, 0.31] |
| 15-year change in Nosos | 0.22** | [0.13, 0.31] | 0.21** | [0.11, 0.30] | 0.19** | [0.09, 0.29] | 0.15** | [0.04, 0.26] |

*Note:* Each model adds covariates to the model, first demographics (sex, race and ethnicity, and education) and technical covariates (chip type, cell proportions), then clinical biomarkers (body mass, blood pressure, and heart rate), and then self-reported smoking and smoking methylation scores. CCI outcomes were estimated with Poisson regression using Monte Carlo simulation to account for missing data; Nosos outcomes were estimated with linear regression using full information maximum likelihood. CI = confidence interval.
\* $p < .05$ . \*\* $p < .01$ .

### Accelerated Aging and Predicted Healthcare Costs

Veterans with faster DunedinPACE aging scores had higher predicted healthcare costs at baseline (β, 0.11; 95% CI, 0.06-0.15, *p* < .001). Veterans with faster aging scores had greater increases in predicted costs over the next 5 years (β, 0.08; 95% CI, 0.03-0.13; *p* < .001), 10 years (β, 0.23; 95% CI, 0.15-0.31; *p* < .001), and 15 years (β, 0.21, 95% CI, 0.11-0.30, *p* < .001). These results largely remained when controlling for clinical biomarkers and smoking measures (**Table 1**). Similar to the CCI, associations increased in size as the follow-up periods increased in length. At the 5-, 10-, and 15-year follow-ups, fastest aging veterans had 11%, 40%, and 38% greater increases in predicted healthcare costs compared to the slowest aging veterans (**eTable 3)**. With estimated annual costs of $14,950 per veteran patient in 202133, these represent $1,645, $5,980, and $5,681 greater increases in annual healthcare expenditures.

### Accelerated Aging and Chronic Disease Incidence

Veterans with faster DunedinPACE aging scores were at increased risk for the onset of any chronic disease comprising the CCI (*HR*, 1.29; 95% CI, 1.21-1.39; *p* < .001). When testing individual chronic diseases individually, faster aging was associated with greater risk for new onset myocardial infarction (84%), stroke (38%), peripheral vascular disease (55%), diabetes (56%), chronic pulmonary disease (19%), cancer (25%), liver disease (44%), and renal disease (34%; **Figure 2**). Results remained when controlling for clinical biomarkers and smoking (**Table 2**), with the exception of peripheral vascular disease and chronic pulmonary disease. **Figure 3** illustrates diabetes onset by DunedinPACE aging scores.

**Figure 2.**
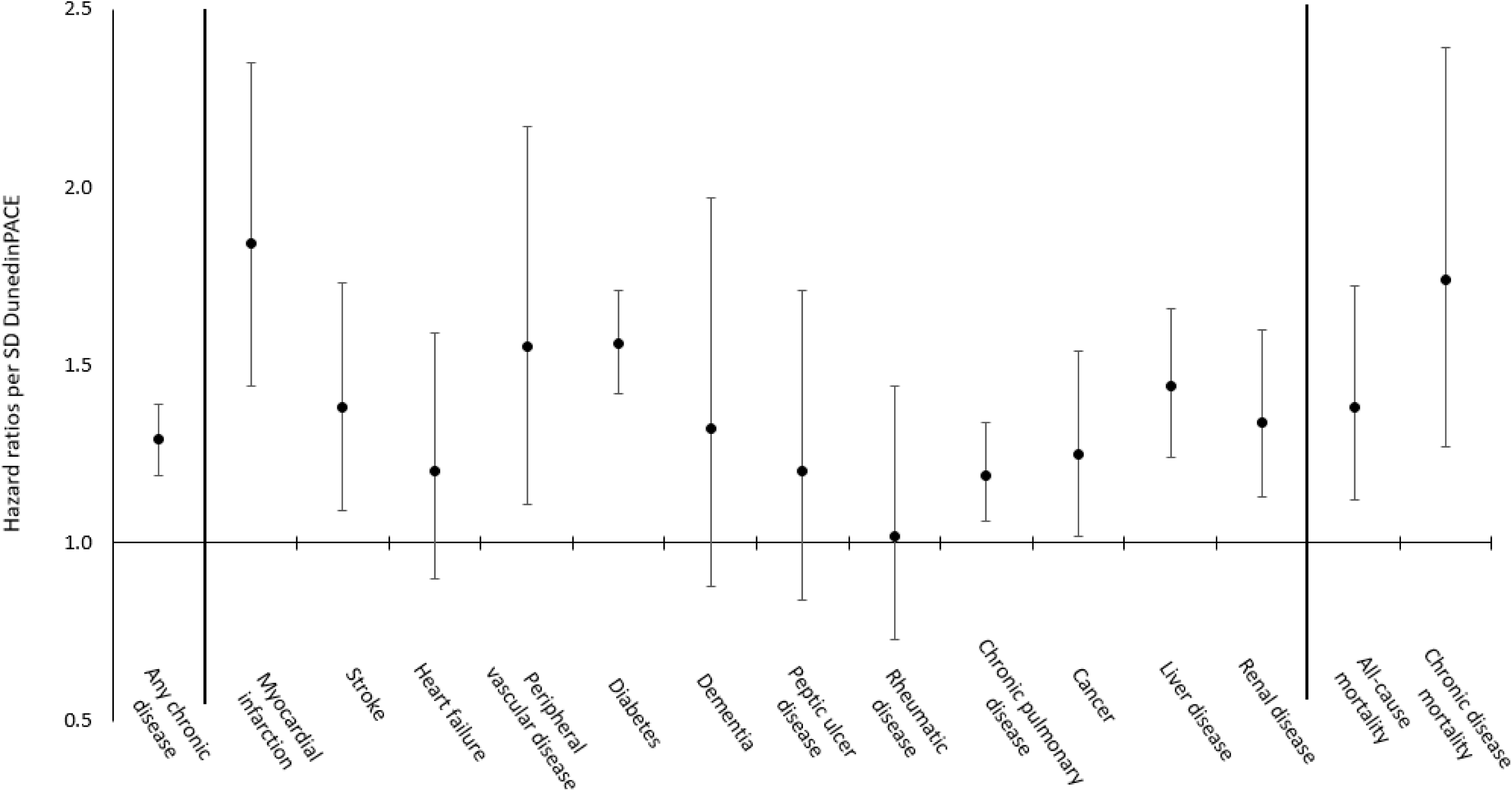
Visualization of the *HR*s for each of the CCI chronic disease categories over the follow-up period. Effects represent *HR*s per 1 *SD* difference in DunedinPACE. All estimates include demographic and technical DNAm covariates. Number of cases and excluded participants for each estimate are presented in **Table 2**. Error bars represent 95% confidence intervals.

**Figure 3.**
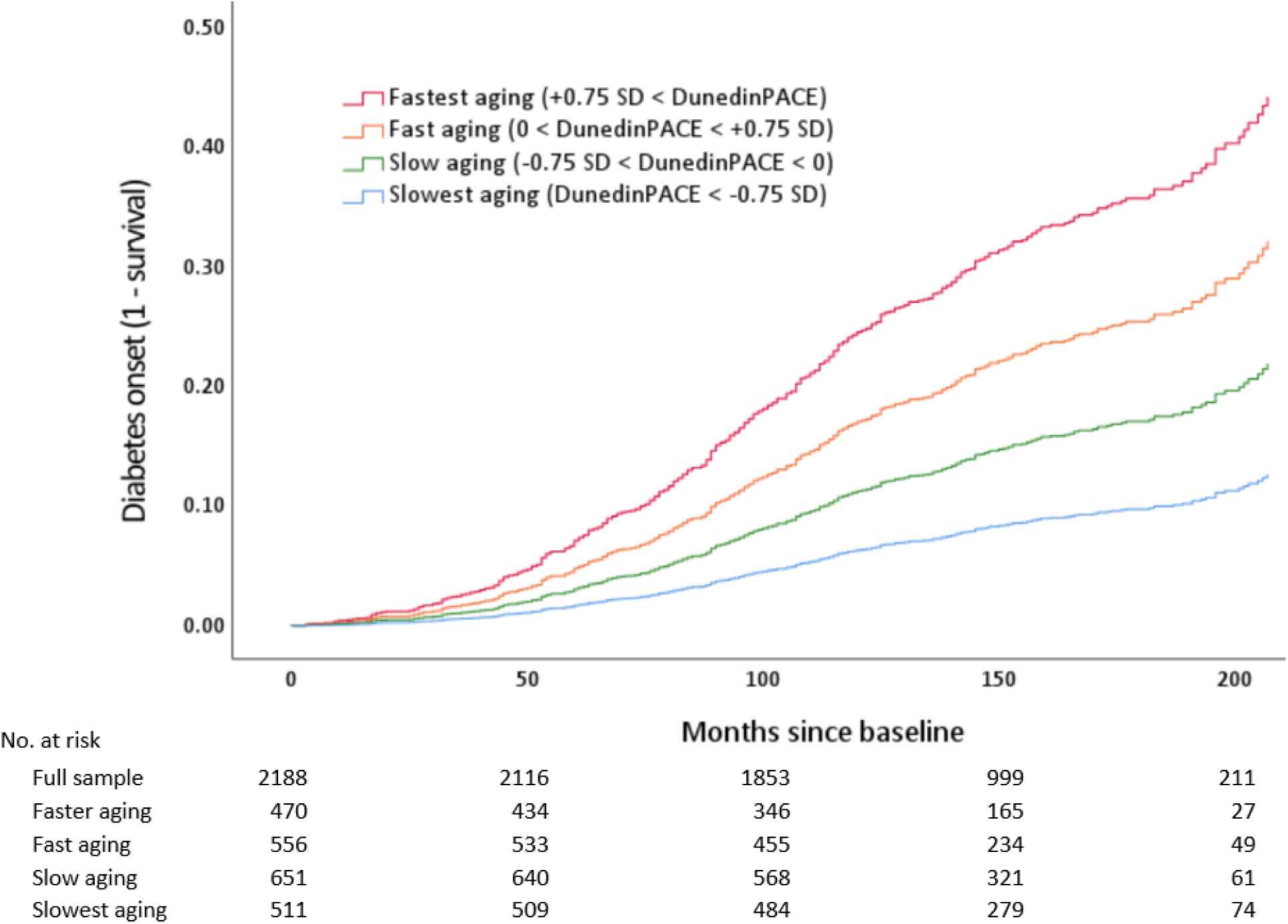
Visualization of diabetes onset (1 – survival) across the follow-up period, as an illustrative example of chronic disease incidence. The model included demographic and technical covariates and excluded 28 veterans with a baseline diagnosis of diabetes or missing covariate data. The four groups were created by standardizing DunedinPACE aging scores and creating cutoffs, as with Figure 1. *No. at risk* represents veterans at risk of diabetes onset at each period on the x-axis. Over the follow up, 45 of 511 (8.8%) slowest aging veterans, 110 of 651 (16.9%) slow aging veterans, 145 of 556 (26.1%) fast aging veterans, and 179 of 470 (38.1%) of fastest aging veterans developed diabetes. Compared to the slowest aging veterans, all other groups were more likely to develop diabetes; slow aging *HR,* 1.90 (95% CI, 1.33-2.70), fast aging *HR,* 3.21 (95% CI, 2.27-4.56), fastest aging *HR*, 5.13 (95% CI, 3.57-7.38, all *p*s < .001).

**Table 2.** Association of DunedinPACE, chronic disease incidence, and mortality among post-9/11 veterans.

| Association with<br>DunedinPACE | Chronic disease at<br>baseline and follow-up |  | Age-adjusted<br>bivariate association |  | Adding demographics<br>and technical covariates |  | Adding clinical<br>biomarkers |  | Adding self-reported and<br>methylation smoking |  |
| --- | --- | --- | --- | --- | --- | --- | --- | --- | --- | --- |
|  | Baseline Dx | Dx Onset | HR | 95% CI | HR | 95% CI | HR | 95% CI | HR | 95% CI |
| **Any chronic disease category** |  |  |  |  |  |  |  |  |  |  |
| Chronic disease onset | *n* = 364 | *n* = 789 | 1.29** | [1.21, 1.39] | 1.28** | [1.19, 1.39] | 1.24** | [1.13, 1.37] | 1.32** | [1.21, 1.45] |
| **Chronic disease categories** |  |  |  |  |  |  |  |  |  |  |
| Myocardial infarction | *n* = 5 | *n* = 56 | 1.64** | [1.32, 2.04] | 1.84** | [1.44, 2.35] | 1.73** | [1.26, 2.37] | 1.61** | [1.17, 2.22] |
| Stroke | *n* = 21 | *n* = 109 | 1.30* | [1.06, 1.61] | 1.38** | [1.09, 1.73] | 1.47** | [1.15, 1.88] | 1.32* | [1.04, 1.68] |
| Heart failure | *n* = 13 | *n* = 53 | 1.38* | [1.07, 1.78] | 1.20 | [0.90, 1.59] | 1.05 | [0.76, 1.44] | 1.07 | [0.77, 1.49] |
| Peripheral vascular disease | *n* = 17 | *n* = 41 | 1.56** | [1.16, 2.08] | 1.55** | [1.11, 2.17] | 1.56* | [1.06, 2.31] | 1.29 | [0.87, 1.90] |
| Diabetes | *n* = 7 | *n* = 482 | 1.54** | [1.41, 1.67] | 1.56** | [1.42, 1.71] | 1.40** | [1.25, 1.57] | 1.85** | [1.66, 2.06] |
| Dementia | *n* = 12 | *n* = 29 | 1.39 | [0.99, 1.97] | 1.32 | [0.88, 1.97] | 1.50 | [0.94, 2.39] | 1.22 | [0.77, 1.94] |
| Peptic ulcer disease | *n* = 17 | *n* = 36 | 1.29 | [0.94, 1.77] | 1.20 | [0.84, 1.71] | 1.09 | [0.73, 1.62] | 1.14 | [0.76, 1.72] |
| Rheumatic disease | *n* = 14 | *n* = 43 | 1.07 | [0.79, 1.45] | 1.02 | [0.73, 1.44] | 1.18 | [0.80, 1.74] | 0.93 | [0.63, 1.38] |
| Chronic pulmonary disease | *n* = 234 | *n* = 325 | 1.20** | [1.07, 1.34] | 1.19** | [1.06, 1.34] | 1.25** | [1.08, 1.44] | 1.08 | [0.94, 1.24] |
| Cancer | *n* = 42 | *n* = 111 | 1.21* | [1.00, 1.45] | 1.25* | [1.02, 1.54] | 1.35* | [1.07, 1.71] | 1.28* | [1.01, 1.63] |
| Liver disease | *n* = 24 | *n* = 206 | 1.35** | [1.18, 1.55] | 1.44** | [1.24, 1.66] | 1.34** | [1.13, 1.58] | 1.52** | [1.27, 1.81] |
| Renal disease | *n* = 26 | *n* = 153 | 1.30** | [1.11, 1.51] | 1.34** | [1.13, 1.60] | 1.36** | [1.11, 1.67] | 1.46** | [1.20, 1.78] |
| **Mortality** |  |  |  |  |  |  |  |  |  |  |
| All-cause mortality | — | *n* = 92 | 1.28* | [1.02, 1.59] | 1.38** | [1.12, 1.72] | 1.25 | [0.97, 1.62] | 1.06 | [0.82, 1.38] |
| Chronic disease mortality | — | *n* = 40 | 1.55** | [1.16, 2.06] | 1.74** | [1.27, 2.39] | 1.59* | [1.10, 2.32] | 1.63** | [1.13, 2.36] |

Note: Each disease specific model excluded participants with the disease at baseline from the cohort *N* of 2,216. Models assessing the inclusion of clinical biomarkers and smoking measures were run with those sets of covariates separately to reduce missing data for the models that included smoking measures. Demographic and technical covariates models also excluded 21 participants who were missing data, clinical biomarker models excluded 561 participants with missing biomarker and education data, and models including self-reported smoking excluded 29 participants missing self-reported smoking or education data. Demographic variables included age, sex, race and ethnicity, education, and technical covariates included chip type and white blood cell proportions. CI = confidence interval; Dx = diagnosis, HR = hazard ratio.
\* $p < .05$ . \*\* $p < .01$ .

### Accelerated Aging and Mortality

Veterans with faster DunedinPACE aging scores were more likely to die due to all causes (*HR,* 1.38; 95% CI 1.12-1.72, *p* = .016). Notably, when excluding mortality due to acute events, the association between aging scores and mortality was approximately twice as strong (*HR*, 1.74, 95% CI 1.27-2.39, *p* < .001). DunedinPACE remained associated with mortality due to chronic disease when also controlling for clinical covariates and smoking (**Table 2**). Aging scores were not associated with mortality due to acute events (*HR,* 0.96, 95% CI 0.63-1.33, *p* = .836).

### Results Stratified by Sex, Race and Ethnicity

We examined associations for CCI and Nosos scores when stratifying by sex, then by race and ethnicity. Men and women veterans showed largely similar associations of DunedinPACE with CCI and Nosos scores at baseline, as well as change over the next 5, 10, and 15 years (**eTable 4)**. Non-Hispanic Black veterans and non-Hispanic White veterans also showed similar associations of DunedinPACE with CCI and Nosos scores at baseline, as well as change over the next 5, 10, and 15 years (**eTable 5)**.

### Results for PC-adjusted Second-generation Epigenetic Clocks

We focused on DunedinPACE epigenetic aging scores, as DunedinPACE currently represents the most widely used third-generation epigenetic measure trained on longitudinal biomarker data. However, we also tested associations for two widely used second-generation epigenetic clocks, adjusted for principle components to improve reliability^12^ (PC-PhenoAge and PC-GrimAge). Consistent with prior studies, the three measures of aging were moderately correlated (.33 ≤ *r* ≤ .52, all *p*s < .001). When accounting for demographic and technical covariates, PC-PhenoAge was not consistently associated with prospective health. In contrast, PC-GrimAge was largely associated with CCI and Nosos scores, as well as a number of specific chronic diseases, all-cause mortality, and chronic disease mortality. Descriptively, the magnitude of most associations for PC-GrimAge were comparable to those for DunedinPACE, though DunedinPACE associations were larger in magnitude when assessing longitudinal change in CCI scores, particularly over longer periods of follow up. Full results for DunedinPACE, PC-PhenoAge, and PC-GrimAge are presented in **eTable 6**.

## Discussion

Veterans with faster DunedinPACE aging scores developed more chronic disease, showed larger increases in predicted healthcare costs, and were at greater risk of premature mortality, as observed in the VA EHR (mean follow-up, 13.1 years). The sizes of these prospective associations appear clinically significant. After ten years, veterans with faster aging developed approximately one additional chronic disease (0.92 points on the CCI), corresponding to a 4.7% greater increase in 10-year predicted mortality risk compared to veterans with slower aging (5.6% versus 0.9% increase, 6.2 times greater relative risk). Veterans with faster aging also had a 40% larger increase in predicted healthcare costs over the next 10 years, representing $5,980 higher annual costs per VA patient compared to veterans with slower aging. In terms of specific chronic disease morbidity and mortality, a 1 *SD* higher DunedinPACE aging score was associated with a 32% increased risk of developing any chronic disease—including increased risk for incident myocardial infarction (61%), stroke (32%), diabetes (85%), cancer (28%), liver disease (52%), and renal disease (46%)—and a 64% increased risk of death due to chronic disease. These associations accounted for numerous covariates—including chronological age, demographic and technical covariates, clinical biomarkers (body mass, BP, and HR), self-reported smoking, and smoking methylation scores. Notably, associations were largely similar for men and women veterans, as well as for non-Hispanic Black veterans and non-Hispanic White veterans.

Our results provide additional support and validation for the use of epigenetic aging measures as surrogate health outcomes in observational studies of health^13^ and randomized control trials^12^ aiming to slow aging. Although prior studies have linked epigenetic aging to a subset of prospective health outcomes^2-3,38-39^, particularly mortality^2-5,14-15^ using research cohorts, none have used EHR data from an integrated healthcare system in a real-world medical setting. By showing that epigenetic aging scores were associated with incident morbidity and mortality, our findings highlight the potential applications of aging biomarkers, such as DunedinPACE, for research and clinical care. As these algorithms are further refined, they can be validated with data from cohorts such as the PDMH with the goal of achieving levels of reliability and validity that could provide predictive utility for patients and clinical providers in the future.

These results have particular relevance to the Veterans Health Administration (VHA). The post-9/11 cohort (currently over 5 million of the 17.9 million living U.S. veterans^39^) is a growing proportion of patients served by the VHA^40^, including greater numbers of women veterans and veterans from racial and ethnic minority groups^41-42^. Our findings show epigenetic aging is associated with prospective health across these demographic groups, suggesting that future uses for epigenetic aging scores would benefit the increasingly diverse patient population utilizing VHA services. Notably, the post-9/11 cohort of veterans is approaching midlife and older age^42^, periods when chronic disease morbidity and mortality become more pronounced. This risk is also an opportunity. The VHA is the largest integrated healthcare system in U.S., and implementing interventions to address risk factors^13,16,43^ for accelerated aging—such as unhealthy behaviors^13^ or PTSD^19^—could delay or prevent the development of ill health for a large number of veterans. If successful, efforts to slow aging using behavioral treatments^44^ or other potentially geroprotective interventions could reduce healthcare costs and, most importantly, prolong veterans’ independence, health, and wellbeing as they grow older. VA clinical trials could also provide data and guidance for implementing interventions in non-VA populations and healthcare systems.

This study has limitations relevant to interpreting our findings. First, the EHR-derived health outcomes can only capture VA clinical encounters or referrals to community care through VA sources. Although it is unlikely that aging and chronic disease coding vary systematically by the amount or type of community care that veterans access through private insurance, it is possible our data are not representative of all veterans’ health. Second, it is not clear to what extent results would generalize to non-veteran populations. It will be important to validate these results in other healthcare systems. Finally, the length of the EHR follow-up varied based on the year that veterans enrolled in the PDMH cohort. Although aging scores were not correlated with enrollment date and results replicated both when accounting for missing data and using listwise deletion, continuing to replicate our findings as the lengths of EHR observation increases in duration would provide additional confidence in our findings.

## Conclusions

Epigenetic aging scores were associated with increased risk for chronic disease morbidity and mortality, as observed in VA medical records for 2,216 U.S veterans^12^ who served after September 11, 2001. Consistent with geroscience theory, faster aging was associated with poorer prospective health across multiple chronic disease categories and organ systems. Epigenetic measures of aging, such as DunedinPACE, might be useful surrogate outcomes for clinical trials and observational cohort studies. With refinement and validation, epigenetic measures of aging might also serve as clinical biomarkers of future health risk that can identify individuals who might be candidates for geroprotective interventions.

## Data Availability

Data from the Post Deployment Mental Health (PDMH) Study are part of a Veterans Affairs data repository and are available to researchers who request access through the VISN 6 MIRECC and follow the appropriate data access protocols. Medical record data from the Veteran Affairs Corporate Data Warehouse are available to researchers who request and are approved for access through the Office of Research and Development (ORD) Data Access Request Tracker (DART). Mortality Data Repository (MDR) data is available to researchers who request MDR data through an MDR Data Request.

## Author access to data

Dr. Kyle Bourassa had full access to all the data in the study and takes responsibility for the integrity of the data and the accuracy of the data analysis.

## Contributions

*Concept and design:* Bourassa, Caspi, Taylor, Hall, Moffitt, Kimbrel. *Acquisition, analysis, or interpretation of data*: Bourassa, Anderson, Woolson, Dennis, Garrett, Sugden, Williams, Houts, Naylor, Ashley-Koch. *Drafting of the manuscript*: Bourassa. *Critical revisions of the manuscript for important intellectual content*: Anderson, Woolson, Dennis, Garrett, Sugden, Williams, Houts, VA Mid Atlantic MIRECC Workgroup, Calhoun, Naylor, Ashley-Koch, Beckham, Caspi, Taylor, Hall, Moffitt, Kimbrel. *Statistical analysis*: Bourassa, Dennis, Garrett, Sugden, Houts, Ashley-Koch. *Obtained funding:* Bourassa, Naylor, Beckham, Kimbrel. *Administrative, technical, or material support:* Garrett, Hair, Dennis. *Supervision*: Taylor, Hall, Moffitt, Kimbrel.

## Disclaimer

The views expressed in this article are those of the authors and do not necessarily reflect the position or policy of the VA, the U.S. government or any other affiliated institution.

## Group information

The VA Mid-Atlantic MIRECC Workgroup contributors include: Eric Dedert, PhD, Eric B. Elbogen, PhD, Robin A. Hurley, MD, Jason D. Kilts, PhD, Angela Kirby, MS, Scott D. McDonald, PhD, Sarah L. Martindale, Ph.D, Christine E. Marx, MD, MS, Scott D. Moore, MD, PhD, Rajendra A. Morey, MD, MS, Jared A. Rowland, PhD, Robert D. Shura, PsyD, Cindy Swinkels, PhD, H. Ryan Wagner, PhD.

## Supplemental Materials

**eFigure 1.**
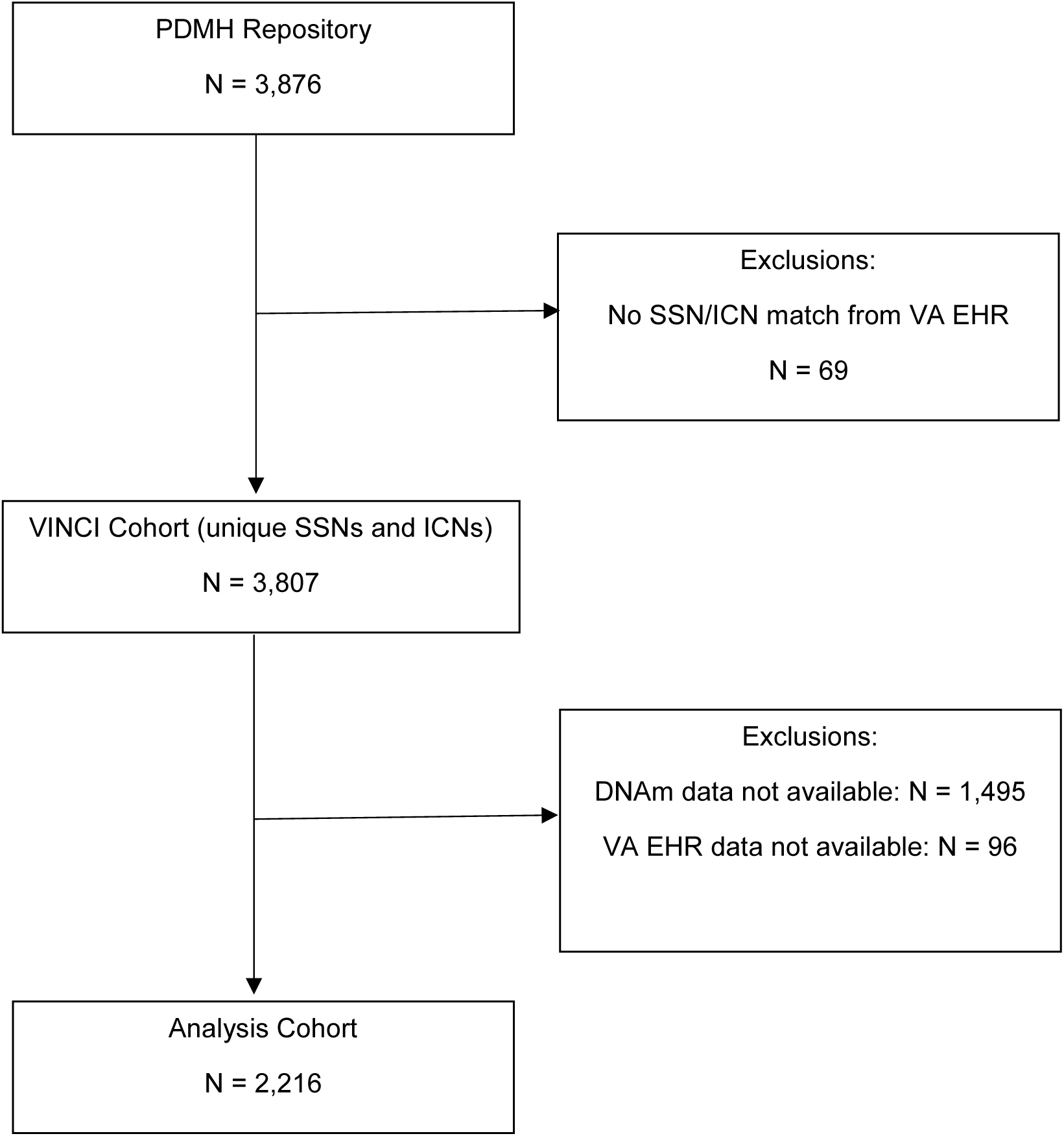
CONSORT-style diagram showing the selection of the final analysis cohort from the original PDMH repository for the current study.

### eMethod 1

#### Detailed description of VA EHR data generation

As described in the main text, prospective health outcomes and clinical biomarkers were derived from data in the VA EHR. Veterans were enrolled in the Post-Deployment Mental Health (PDMH^1^) study with a baseline that ranged from 2005 to 2016, which resulted in follow-up periods that ranged from 7.3 to 18.5 years (88 to 222 months; **eFigure 2**). After project approval through the Durham VA Research & Development committee and VA Data Access Request Tracker (DART), data linkage was completed in the VA Informatics and Computing Infrastructure (VINCI) system using social security numbers (SSNs) from the PDMH linked to VA patient’s SSN and internal control number (ICN). Data for the relevant health outcomes were then called from VA Corporate Data Warehouse (CDW) tables using SQL coding. Chronic disease and biomarker data were generated from outpatient, inpatient, and purchased care data (e.g., community care that is delivered to veterans following referral from the VA and/or paid by VA sources). Data cleaning processes are reported in the following sections. EHR data coverage and missingness based on length of follow-up is provided in **eTable 1.** Data pulls were completed during early 2024, with a final refresh of CDW data for the current study on 5/15/2024.

#### Charlson comorbidity index

Charlson comorbidity index scores were derived using diagnostic ICD-9 and ICD-10 codes^2^, matching the tables crated by Glasheen and colleagues. Baseline values used ICD codes at the date of enrollment in the PDMH and were updated for each subsequent year. Chronic diseases diagnoses were carried forward to subsequent years. Scores were coded as missing for any year past the end of the veteran’s follow-up observation period, as well as if there was insufficient evidence of a patient utilizing VA healthcare and no diagnoses were present.

#### Nosos risk adjustment score

Nosos risk adjustment scores^3^ are automatically calculated in the VA CDW. Missing scores were due to either the length of follow-up or the missingness in the VA CDW. Nosos scores reflected values for the algorithm that did not account for pharmacy costs.

#### Chronic disease diagnoses

As described in the main text, chronic disease diagnostic statuses were ascertained by presence of ICD codes in veterans’ EHR at baseline, as well at follow-up to a censor date of 12/31/2023. As with the CCI, ICD-9 and ICD-10 codes were drawn directly from tables outlined by Glasheen and colleagues^1^. For disease categories with multiple severities (diabetes, liver disease, renal disease, and cancer), we used earliest onset of any severity. HIV/AIDS and hemiplegia were not investigated individually due to low incidence over the follow-up (*n* < 20), but remained in overall CCI scores.

#### Clinic-assessed biomarkers

Clinical biomarkers assessed in VA encounters included body mass, blood pressure (BP), and heart rate (HR). Body mass was calculated using the standard formula utilizing height and weight. All measures were assessed at baseline using an average of the three valid scores closest to the PDMH baseline up to a 2-year lookback period. Any height measurements lower than 61cm and higher than 305 cm, or more than 25 cm from a veteran’s mean, were excluded. Any weight measurements less than 4.5 kg and more than 454 kg were excluded. Weight changes defined by a criteria of percent change relative to nearest neighbor relative to overall mean, and time from nearest neighbor (i.e., change greater than 10% and more than 1 *SD* from the mean in one week) were excluded^4^. BP was assessed using systolic BP. Systolic BP measurements less than 50 mmHg were excluded. HR was assessed using pulse. HR measurements below 30 and more than 200 beats/minute were excluded.

#### Mortality

All-cause mortality status was ascertained using dates of death pulled from the VA CDW’s death ascertainment file and confirmed with National Death Index (NDI) through the VA Mortality Data Repository^5^ (MDR). VA CDW data were censored to a date of 1/21/2024, whereas MDR data are currently available until 12/31/2021^4^. Acute versus chronic disease mortality was assessed using the primary ICD code associated with the mortality event.

---

**eTable 1.** Coverage for study variables.

| **Overall sample N = 2,216** | **Total n** | **% of total N** |
| --- | --- | --- |
| **Charlson comorbidity index scores** |  |  |
| Baseline CCI score | 2,216 | 100% |
| 5-year change in CCI | 2,216 | 100% |
| 10-year change in CCI | 1,889 | 85.2% |
| 15-year change in CCI | 624 | 28.2% |
| **Nosos scores** |  |  |
| Baseline Nosos score | 1,827 | 82.4% |
| 5-year change in Nosos | 1,641 | 74.1% |
| 10-year change in Nosos | 1,284 | 57.9% |
| 15-year change in Nosos | 315 | 14.2% |
| **Clinical biomarkers** |  |  |
| Body mass | 1,676 | 75.6% |
| Blood pressure | 1,756 | 79.2% |
| Heart rate | 1,753 | 79.1% |
| **Demographic characteristics** |  |  |
| Age | 2,216 | 100% |
| Sex | 2,216 | 100% |
| Race and ethnicity | 2,216 | 100% |
| Education | 2,195 | 99.1% |
| Self-reported smoking | 2,206 | 99.5% |

*Note:* All measures derived from DNA methylation (aging scores, technical covariates, and methylation smoking scores) had full data for all participants.

**eFigure 2.**
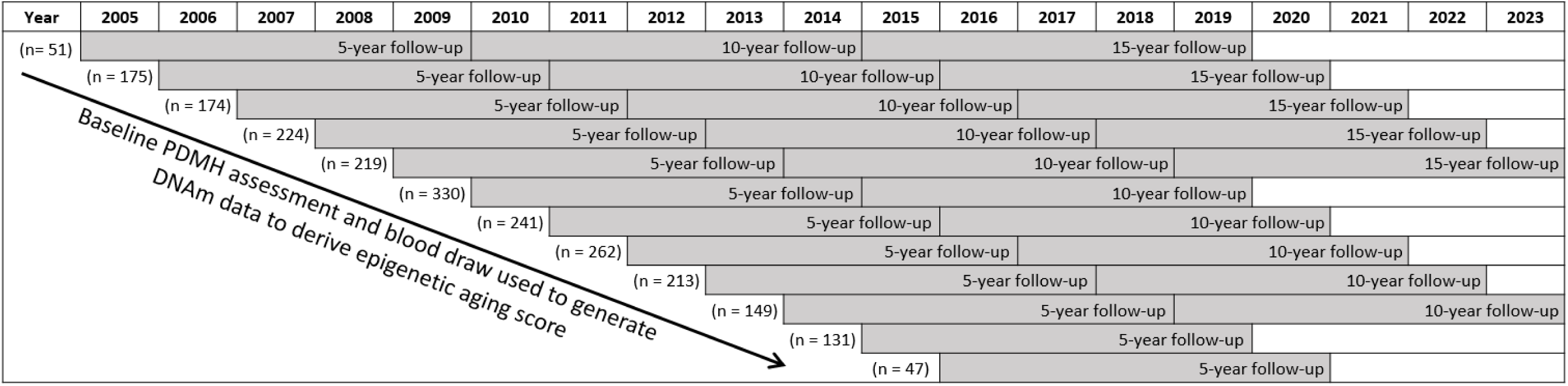
Illustration of cohort enrollment and follow-up timelines by year of baseline PDMH participation. Follow-ups represent veteran data used for models testing associations for change in CCI and Nosos scores to the 5-, 10-, and 15-year follow-ups. Models testing associations of DunedinPACE with chronic disease incidence and mortality used all follow-up data.

**eFigure 3.**
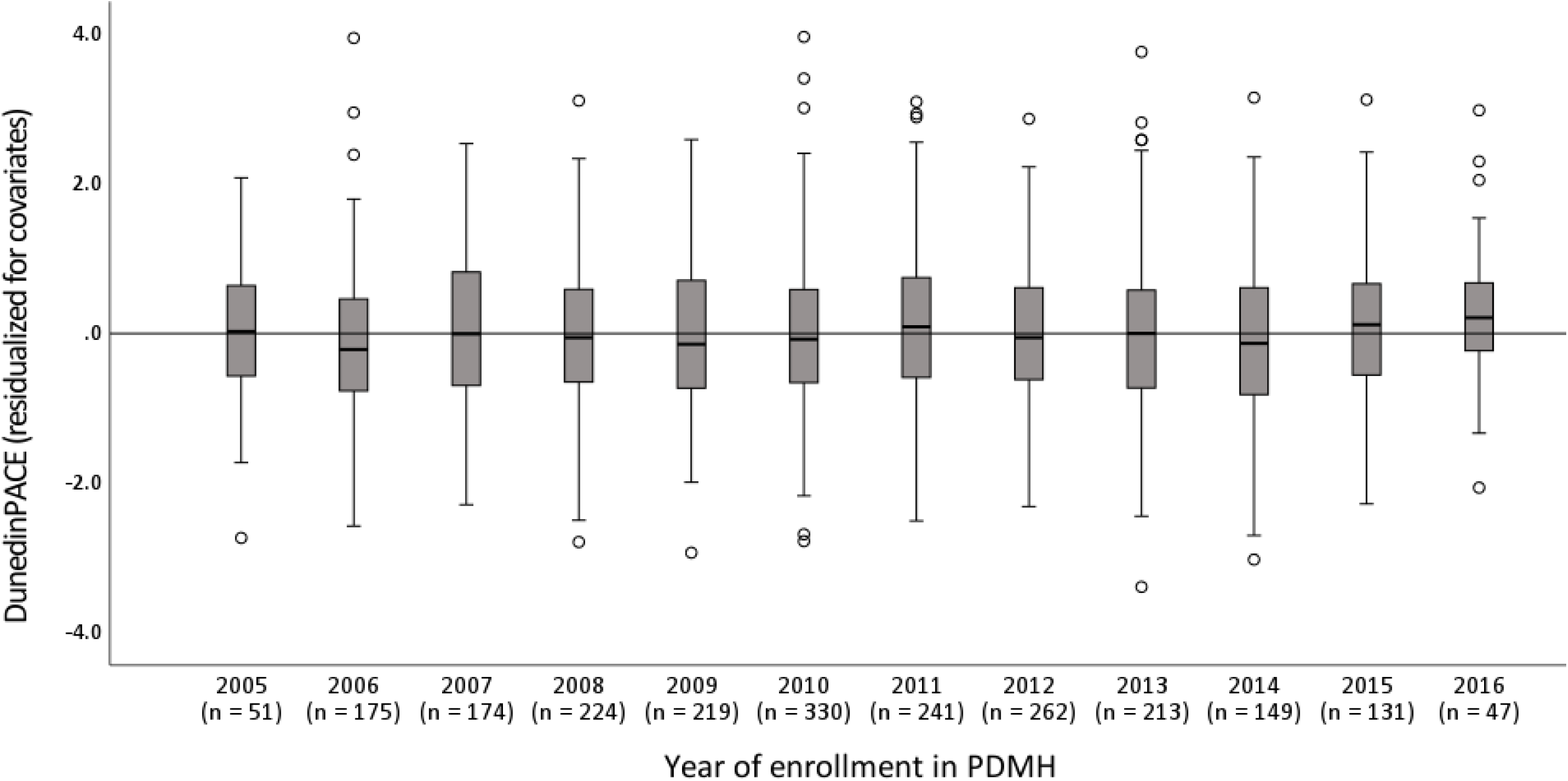
DunedinPACE scores (residualized for age, demographic covariates, and technical covariates) organized by year of baseline assessment in the PDMH. There was not a significant association between year of enrollment (which accounted for years of follow-up) and DunedinPACE aging scores (*r*, .02; 95% CI, -.02-.07; *p* = .256).

**Figure S4.**
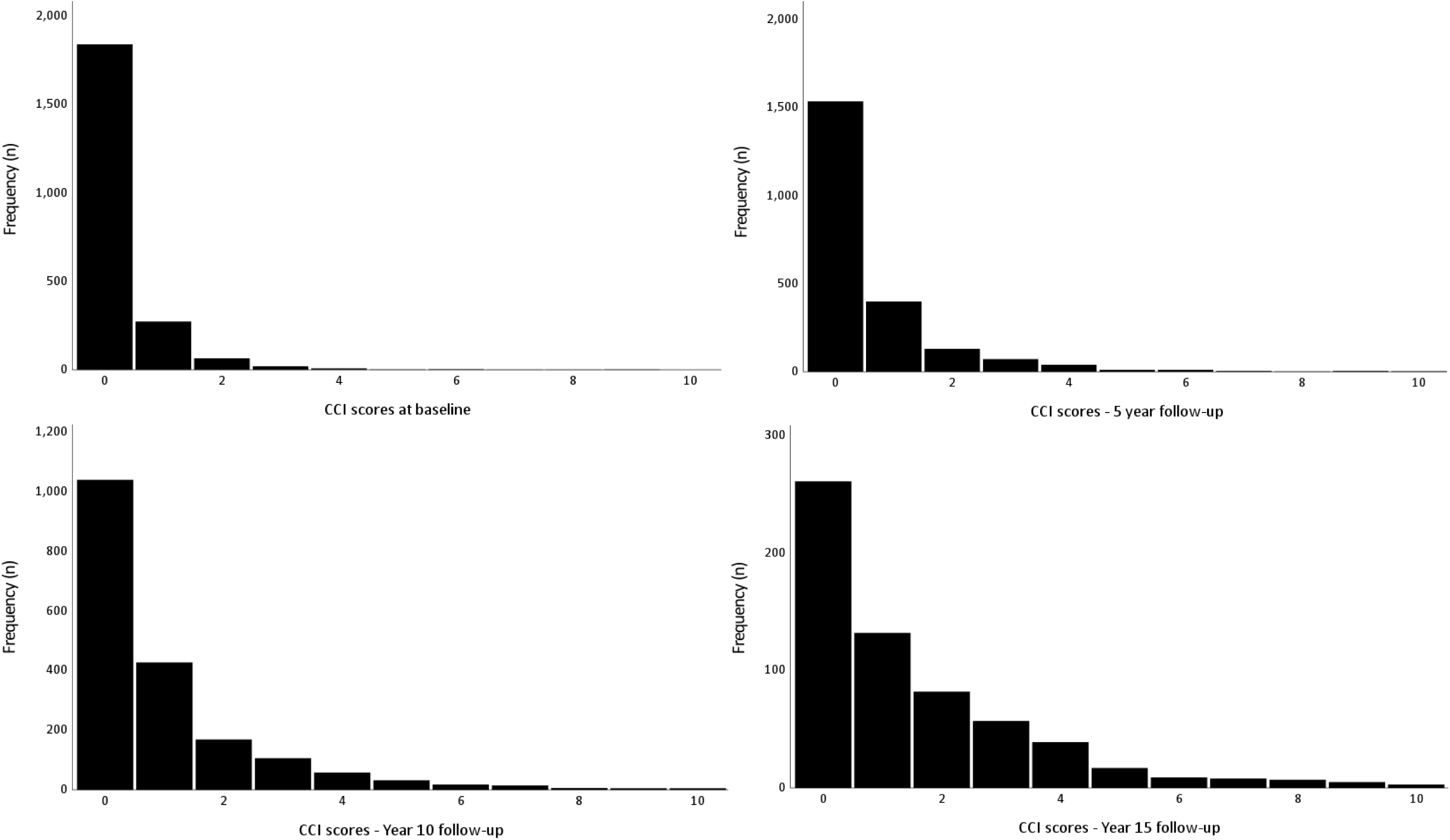
Distributions of CCI scores at baseline and the 5-, 10-, and 15-year follow-up. Total *N*s reflect overall observation periods due to differences in baseline year of assessment (Baseline *n,* 2,216; 5-year *n,* 2,216; 10-year *n,* 1,889; 15-year *n*, 624).

**eTable 2.** Study results from models with CCI and Nosos scores excluding participants with missing data.

| Association with<br>DunedinPACE | Age-adjusted<br>bivariate association |  | Adding demographic and<br>technical covariates |  | Adding clinical<br>biomarkers |  | Adding self-reported and<br>methylation smoking |  |
| --- | --- | --- | --- | --- | --- | --- | --- | --- |
| | $\beta$ | 95% CI | $\beta$ | 95% CI | $\beta$ | 95% CI | $\beta$ | 95% CI |
| **Chronic disease burden (Charlson comorbidity index score)** |  |  |  |  |  |  |  |  |
| Baseline CCI score | 0.25** | [0.14, 0.36] | 0.22** | [0.09, 0.34] | 0.20** | [0.07, 0.34] | 0.23** | [0.08, 0.38] |
| 5-year change in CCI | 0.20** | [0.12, 0.28] | 0.21** | [0.12, 0.30] | 0.16** | [0.06, 0.25] | 0.12* | [0.01, 0.22] |
| 10-year change in CCI | 0.19** | [0.12, 0.26] | 0.24** | [0.16, 0.31] | 0.19** | [0.11, 0.27] | 0.17** | [0.08, 0.25] |
| 15-year change in CCI | 0.29** | [0.18, 0.37] | 0.31** | [0.20, 0.42] | 0.24** | [0.13, 0.36] | 0.29** | [0.17, 0.41] |
| **Predicted annual VA healthcare costs (Nosos risk adjustment score)** |  |  |  |  |  |  |  |  |
| Baseline Nosos score | 0.10** | [0.05, 0.14] | 0.12** | [0.07, 0.17] | 0.12** | [0.07, 0.19] | 0.05 | [-0.02, 0.12] |
| 5-year change in Nosos | 0.08** | [0.03, 0.13] | 0.07* | [0.01, 0.13] | 0.06 | [-0.00, 0.12] | 0.02 | [-0.05, 0.09] |
| 10-year change in Nosos | 0.18** | [0.12, 0.22] | 0.17** | [0.10, 0.22] | 0.18** | [0.10, 0.24] | 0.15** | [0.07, 0.23] |
| 15-year change in Nosos | 0.23** | [0.10, 0.30] | 0.20** | [0.06, 0.29] | 0.18* | [0.03, 0.28] | 0.16* | [0.00, 0.28] |

*Note:* All models exclude participants with missing data. Each model adds more covariates to the model, first demographics (sex, race and ethnicity, and education) and technical covariates (chip type, cell proportions), then clinical biomarkers (body mass, blood pressure, and heart rate), and then self-reported smoking and smoking methylation scores. CCI outcomes were estimated with Poisson regression, Nosos outcomes were estimated with linear regression. CI = confidence interval.
\* $p < .05$ . \*\* $p < .01$ .

**Figure S5.**
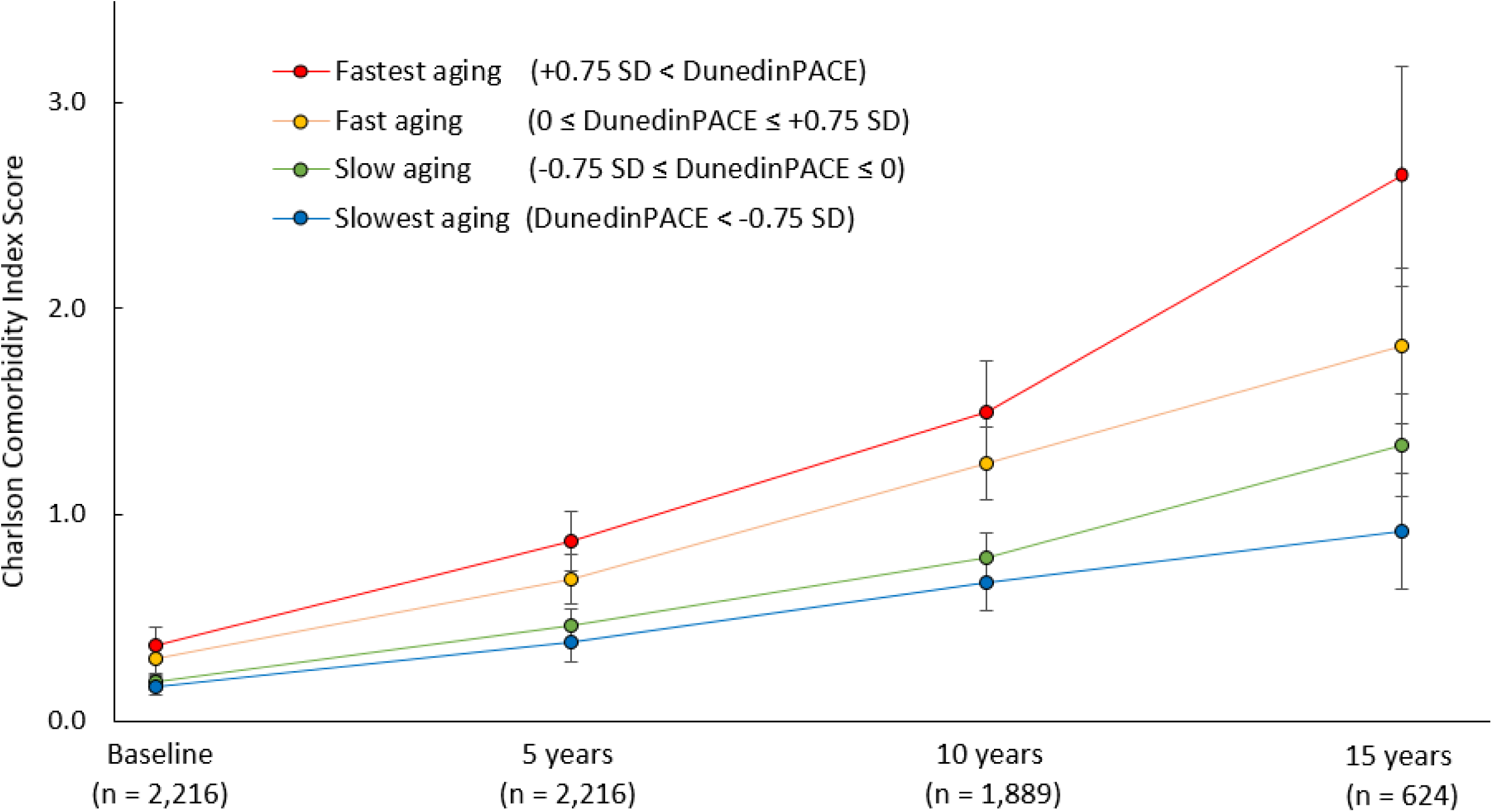
CCI scores over time grouped by DunedinPACE scores that were residualized for age. Four groups were created by standardizing DunedinPACE aging scores residualized for age and creating cutoffs at the mean and 0.75 *SD* above and below the mean, “slowest aging” (*n* = 504, 22.7%), “slow aging” (*n* = 681, 30.7%), “fast aging” (*n* = 551, 24.9%), and “fastest aging” (*n* = 480, 21.7%).

**eTable 3.** 10-year CCI-predicted mortality by DunedinPACE aging groups at baseline, and 5-, 10-, and 15-year follow-ups.

|  | Baseline | 5-year follow-up |  | 10-year follow-up |  | 15-year follow-up |  |
| --- | --- | --- | --- | --- | --- | --- | --- |
| 10-year predicted mortality risk | Mortality risk | Mortality risk | Change in risk | Mortality risk | Change in risk | Mortality risk | Change in risk |
| Slowest aging | 1.9% | 2.2% | 0.3% | 2.8% | 0.9% | 3.4% | 1.5% |
| Slow aging | 2.0% | 2.5% | 0.5% | 3.3% | 1.4% | 5.5% | 3.5% |
| Fast aging | 2.2% | 3.1% | 0.9% | 4.7% | 2.4% | 8.4% | 6.2% |
| Fastest aging | 3.4% | 4.1% | 1.6% | 8.1% | 5.6% | 20.5% | 18.0% |
| Annual predicted VA healthcare costs | Nosos score | Nosos score | Change in score | Nosos score | Change in score | Nosos score | Change in score |
| Slowest aging | 0.73 | 0.80 | 7% | 0.77 | 4% | 0.84 | 11% |
| Slow aging | 0.80 | 0.87 | 7% | 0.89 | 9% | 0.75 | -5% |
| Fast aging | 0.91 | 0.95 | 4% | 0.98 | 7% | 1.02 | 11% |
| Fastest aging | 0.92 | 1.10 | 18% | 1.36 | 44% | 1.42 | 49% |

*Note:* All results represent means generated for each group from raw values for the participants with data at that occasion. 10-year CCI-predicted mortality risk values were calculated using the standard calculations for the CCI risk index [Percent mortality risk = $100 \times (1 - (0.983^{e^{CCI}} \times 0.9))$ ] applied to the mean values for each group. Change in score columns represent changes from baseline values to each follow-up. Participant *Ns* match those reported for the CCI and Nosos scores at each time point in **eTable 1**; CCI at baseline *n*, 2,216; 5-year *n*, 2,216; 10-year *n*, 1,889; 15-year *n*, 624; Nosos *n*, 1,827; 5-year *n*, 1,641; 10-year *n*, 1,284; 15-year *n*, 315. Groups were created by standardizing DunedinPACE scores and dividing veterans into four groups with cutoffs at the mean, as well as *SD* = 0.75 above and below the mean. “Slowest aging” represented values more than -0.75 *SD* below the mean, “slow aging” represented values from -0.75 to 0, “fast aging” represented values from 0 to +0.75 *SD*, and “fastest aging” represented values more than +0.75 *SD* above the mean. These groups were created using *a priori* cutoffs for illustrative purposes only—all model estimates for CCI and Nosos scores reported in the paper were conducted using continuous outcomes.

**eTable 4.** Association of DunedinPACE and health stratified by sex.

| Association with<br>DunedinPACE | Age-adjusted<br>bivariate association |  | Adding demographic and<br>technical covariates |  | Adding clinical<br>biomarkers |  | Adding self-reported and<br>methylation smoking |  |
| --- | --- | --- | --- | --- | --- | --- | --- | --- |
| N = 2,216 | $\beta$ | 95% CI | $\beta$ | 95% CI | $\beta$ | 95% CI | $\beta$ | 95% CI |
| **Baseline CCI score** |  |  |  |  |  |  |  |  |
| Men veterans | 0.24** | [0.10, 0.38] | 0.22** | [0.07, 0.37] | 0.17 | [0.00, 0.34] | 0.13 | [-0.10, 0.37] |
| Women veterans | 0.15* | [0.01, 0.29] | 0.14 | [-0.02, 0.29] | 0.16 | [-0.01, 0.32] | 0.22* | [0.03, 0.41] |
| **5-year change in CCI score** |  |  |  |  |  |  |  |  |
| Men veterans | 0.19** | [0.10, 0.28] | 0.18** | [0.08, 0.29] | 0.13* | [0.02, 0.24] | 0.11 | [-0.00, 0.24] |
| Women veterans | 0.14 | [-0.01, 0.29] | 0.20* | [0.04, 0.36] | 0.16 | [-0.07, 0.39] | 0.14 | [-0.04, 0.31] |
| **10-year change in CCI score** |  |  |  |  |  |  |  |  |
| Men veterans | 0.18** | [0.11, 0.26] | 0.24** | [0.15, 0.33] | 0.21** | [0.12, 0.30] | 0.20** | [0.10, 0.30] |
| Women veterans | 0.19* | [0.03, 0.34] | 0.22** | [0.06, 0.39] | 0.14 | [-0.09, 0.55] | 0.16 | [-0.07, 0.39] |
| **15-year change in CCI score** |  |  |  |  |  |  |  |  |
| Men veterans | 0.26** | [0.18, 0.34] | 0.30** | [0.17, 0.43] | 0.27** | [0.14, 0.40] | 0.30** | [0.16, 0.43] |
| Women veterans | 0.38** | [0.17, 0.60] | 0.39** | [0.11, 0.67] | 0.38* | [0.08, 0.68] | 0.37* | [0.05, 0.69] |
| **Baseline Nosos score** |  |  |  |  |  |  |  |  |
| Men veterans | 0.09** | [0.04, 0.14] | 0.11** | [0.06, 0.17] | 0.11** | [0.06, 0.17] | 0.05 | [-0.01, 0.11] |
| Women veterans | 0.07 | [-0.01, 0.14] | 0.09* | [0.01, 0.17] | 0.09* | [0.00, 0.18] | 0.04 | [-0.05, 0.14] |
| **5-year change in Nosos score** |  |  |  |  |  |  |  |  |
| Men veterans | 0.10** | [0.05, 0.16] | 0.09** | [0.02, 0.15] | 0.08* | [0.01, 0.14] | 0.04 | [-0.03, 0.12] |
| Women veterans | 0.08* | [0.00, 0.15] | 0.06 | [-0.02, 0.14] | 0.04 | [-0.04, 0.13] | 0.01 | [-0.08, 0.10] |
| **10-year change in Nosos score** |  |  |  |  |  |  |  |  |
| Men veterans | 0.21** | [0.14, 0.28] | 0.21** | [0.13, 0.29] | 0.20** | [0.11, 0.28] | 0.17** | [0.07, 0.26] |
| Women veterans | 0.32** | [0.13, 0.52] | 0.26* | [0.05, 0.48] | 0.30* | [0.07, 0.53] | 0.31* | [0.06, 0.56] |
| **15-year change in Nosos score** |  |  |  |  |  |  |  |  |
| Men veterans | 0.24** | [0.14, 0.35] | 0.24** | [0.12, 0.35] | 0.20** | [0.09, 0.32] | 0.12 | [-0.01, 0.25] |
| Women veterans | 0.17* | [0.01, 0.34] | 0.10 | [-0.04, 0.24] | 0.13 | [-0.03, 0.28] | 0.16 | [-0.00, 0.33] |

*Note:* Each model adds more covariates to the model, first demographics (race and ethnicity, and education) and technical covariates (chip type, cell proportions), then clinical biomarkers (body mass, blood pressure, and heart rate), and then self-reported smoking and smoking methylation scores. CCI outcomes were estimated with Poisson regression using Monte Carlo simulation to account for missing data; Nosos outcomes were estimated with linear regression using full information maximum likelihood. CI = confidence interval.
\* $p < .05$ . \*\* $p < .01$ .

**eTable 5.** Association of DunedinPACE and health stratified by race and ethnicity.

| Association with<br>DunedinPACE<br>N = 2,216 | Age-adjusted<br>bivariate association |  | Adding demographic and<br>technical covariates |  | Adding clinical<br>biomarkers |  | Adding self-reported and<br>methylation smoking |  |
| --- | --- | --- | --- | --- | --- | --- | --- | --- |
| | $\beta$ | 95% CI | $\beta$ | 95% CI | $\beta$ | 95% CI | $\beta$ | 95% CI |
| **Baseline CCI score** |  |  |  |  |  |  |  |  |
| Non-Hispanic Black veterans | 0.21* | [0.05, 0.37] | 0.17* | [0.01, 0.34] | 0.17* | [0.01, 0.34] | 0.21* | [0.02, 0.40] |
| Non-Hispanic White veterans | 0.27** | [0.05, 0.17] | 0.29** | [0.09, 0.48] | 0.22 | [-0.01, 0.45] | 0.12 | [0.20, 0.44] |
| **5-year change in CCI score** |  |  |  |  |  |  |  |  |
| Non-Hispanic Black veterans | 0.20** | [0.09, 0.30] | 0.20** | [0.07, 0.32] | 0.13* | [0.01, 0.26] | 0.14* | [0.01, 0.26] |
| Non-Hispanic White veterans | 0.18** | [0.06, 0.31] | 0.24** | [0.11, 0.36] | 0.18* | [0.04, 0.33] | 0.22* | [0.03, 0.40] |
| **10-year change in CCI score** |  |  |  |  |  |  |  |  |
| Non-Hispanic Black veterans | 0.22** | [0.11, 0.32] | 0.27** | [0.15, 0.37] | 0.19** | [0.08, 0.29] | 0.20** | [0.09, 0.31] |
| Non-Hispanic White veterans | 0.17** | [0.07, 0.28] | 0.25** | [0.14, 0.37] | 0.17** | [0.04, 0.30] | 0.14 | [-0.01, 0.29] |
| **15-year change in CCI score** |  |  |  |  |  |  |  |  |
| Non-Hispanic Black veterans | 0.27** | [0.12, 0.42] | 0.27** | [0.09, 0.46] | 0.24** | [0.07, 0.41] | 0.32** | [0.15, 0.48] |
| Non-Hispanic White veterans | 0.30** | [0.16, 0.43] | 0.37** | [0.23, 0.52] | 0.32** | [0.18, 0.47] | 0.32** | [0.14, 0.50] |
| **Baseline Nosos score** |  |  |  |  |  |  |  |  |
| Non-Hispanic Black veterans | 0.09** | [0.02, 0.11] | 0.09** | [0.03, 0.14] | 0.08** | [0.04, 0.19] | 0.03 | [-0.03, 0.09] |
| Non-Hispanic White veterans | 0.11** | [0.03, 0.16] | 0.13** | [0.06, 0.20] | 0.13** | [0.06, 0.20] | 0.06 | [-0.03, 0.15] |
| **5-year change in Nosos score** |  |  |  |  |  |  |  |  |
| Non-Hispanic Black veterans | 0.06* | [0.00, 0.12] | 0.05 | [-0.02, 0.11] | 0.03 | [-0.04, 0.09] | 0.02 | [-0.06, 0.09] |
| Non-Hispanic White veterans | 0.13** | [0.05, 0.20] | 0.11** | [0.03, 0.19] | 0.11* | [0.03, 0.20] | 0.06 | [-0.04, 0.16] |
| **10-year change in Nosos score** |  |  |  |  |  |  |  |  |
| Non-Hispanic Black veterans | 0.32** | [0.21, 0.44] | 0.32** | [0.20, 0.45] | 0.32** | [0.19, 0.45] | 0.33** | [0.18, 0.47] |
| Non-Hispanic White veterans | 0.14** | [0.06, 0.22] | 0.14** | [0.06, 0.22] | 0.12** | [0.04, 0.22] | 0.07 | [-0.04, 0.17] |
| **15-year change in Nosos score** |  |  |  |  |  |  |  |  |
| Non-Hispanic Black veterans | 0.19** | [0.06, 0.32] | 0.16* | [0.01, 0.31] | 0.12 | [-0.04, 0.27] | 0.05 | [-0.11, 0.22] |
| Non-Hispanic White veterans | 0.20** | [0.10, 0.35] | 0.16* | [0.03, 0.29] | 0.14* | [0.00, 0.28] | 0.10 | [-0.05, 0.25] |

Note: Each model adds more covariates to the model, first demographics (sex and education) and technical covariates (chip type, cell proportions), then clinical biomarkers (body mass, blood pressure, and heart rate), and then self-reported smoking and smoking methylation scores. CI = confidence interval.
\* $p < .05$ . \*\* $p < .01$ .

**eTable 6.** Association of PC-adjusted second-generation epigenetic clocks and veteran health.

| *N* = 2,216 | Associations with<br>DunedinPACE |  | Associations with<br>PC-GrimAge |  | Associations with<br>PC-PhenoAge |  |
| --- | --- | --- | --- | --- | --- | --- |
| | $\beta$ | 95% CI | $\beta$ | 95% CI | $\beta$ | 95% CI |
| **Chronic disease burden (Charlson comorbidity index score)** |  |  |  |  |  |  |
| Baseline CCI score | 0.23** | [0.10, 0.35] | 0.22** | [0.08, 0.36] | 0.02 | [-0.02, 0.08] |
| 5-year change in CCI | 0.22** | [0.13, 0.31] | 0.15** | [0.06, 0.25] | 0.02 | [-0.07, 0.11] |
| 10-year change in CCI | 0.24** | [0.16, 0.31] | 0.16** | [0.08, 0.24] | 0.03 | [-0.06, 0.12] |
| 15-year change in CCI | 0.31** | [0.20, 0.42] | 0.10 | [-0.01, 0.21] | -0.09 | [-0.22, 0.03] |
| **Predicted annual VA healthcare costs (Nosos risk adjustment score)** |  |  |  |  |  |  |
| Baseline Nosos score | 0.11** | [0.06, 0.15] | 0.16** | [0.11, 0.21] | 0.00 | [-0.04, 0.05] |
| 5-year change in Nosos | 0.08** | [0.03, 0.13] | 0.10** | [0.04, 0.15] | -0.01 | [-0.07, 0.05] |
| 10-year change in Nosos | 0.23** | [0.15, 0.31] | 0.19** | [0.10, 0.27] | 0.09* | [0.01, 0.17] |
| 15-year change in Nosos | 0.21** | [0.11, 0.30] | 0.18** | [0.08, 0.28] | 0.06 | [-0.05, 0.16] |
| **Any chronic disease category** |  |  |  |  |  |  |
| Chronic disease onset | 1.28** | [1.19, 1.39] | 1.18** | [1.08, 1.28] | 1.12** | [1.03, 1.21] |
| **Chronic disease categories** |  |  |  |  |  |  |
| Myocardial infarction | 1.84** | [1.44, 2.35] | 1.75** | [1.36, 2.27] | 1.17 | [0.87, 1.57] |
| Stroke | 1.38** | [1.09, 1.73] | 1.50** | [1.23, 1.85] | 1.03 | [0.74, 1.42] |
| Heart failure | 1.20 | [0.90, 1.59] | 1.24 | [0.92, 1.67] | 1.14 | [0.91, 1.42] |
| Peripheral vascular disease | 1.55** | [1.11, 2.17] | 1.77** | [1.30, 2.41] | 1.20 | [0.84, 1.70] |
| Diabetes | 1.56** | [1.42, 1.71] | 1.14* | [1.03, 1.26] | 1.16** | [1.05, 1.29] |
| Dementia | 1.32 | [0.88, 1.97] | 1.28 | [0.86, 1.92] | 0.90 | [0.58, 1.38] |
| Peptic ulcer disease | 1.20 | [0.84, 1.71] | 1.13 | [0.77, 1.64] | 0.97 | [0.68, 1.38] |
| Rheumatic disease | 1.02 | [0.73, 1.44] | 1.10 | [0.77, 1.59] | 1.02 | [0.73, 1.44] |
| Chronic pulmonary disease | 1.19** | [1.06, 1.34] | 1.27** | [1.13, 1.43] | 1.06 | [0.92, 1.20] |
| Cancer | 1.25* | [1.02, 1.54] | 1.23 | [0.99, 1.52] | 0.91 | [0.73, 1.13] |
| Liver disease | 1.44** | [1.24, 1.66] | 1.21* | [1.04, 1.41] | 1.16 | [0.99, 1.36] |
| Renal disease | 1.34** | [1.13, 1.60] | 1.05 | [0.87, 1.27] | 1.06 | [0.87, 1.28] |
| **Mortality** |  |  |  |  |  |  |
| All-cause mortality | 1.38** | [1.12, 1.72] | 1.53** | [1.26, 1.86] | 1.16 | [0.92, 1.46] |
| Chronic disease mortality | 1.74** | [1.27, 2.39] | 1.69** | [1.26, 2.27] | 1.36 | [0.98, 1.88] |

*Note:* All models included demographic (sex, race and ethnicity, and education) and technical covariates (chip type, cell proportions). CCI outcomes were estimated with Poisson regression, Nosos outcomes were estimated with linear regression, chronic disease and mortality outcomes used Cox proportional-hazards models. CI = confidence interval.
\* $p < .05$ . \*\* $p < .01$ .

